# Burden and bacteriological profile of sepsis among adult medical emergencies presenting to a national referral hospital in Kampala, Uganda

**DOI:** 10.1101/2022.04.26.22274311

**Authors:** Sharon Nyesiga, Jane Nakibuuka, Henry Kajumbula, Ronald Ssenyonga, Pauline Byakika-Kibwika

**Affiliations:** Department of Internal Medicine, Makerere University College of Health Sciences; Department of Critical care Medicine, Mulago National Referral Hospital; Department of Medical Microbiology, Makerere University College of Health Sciences; School of Public Health, Makerere University College of Health Sciences

**Keywords:** Sepsis, anti-microbial resistance, mortality, Uganda

## Abstract

**Introduction:** Sepsis, defined as life-threatening organ dysfunction due to dysregulated host response to infection can result from any infection. In 2017, an estimated 48.9 million incident cases of sepsis and 11.8 million sepsis-related deaths were reported globally. Bacterial infection is the major cause of sepsis. Data about sepsis burden is derived almost exclusively from studies in high-income countries yet mortality from sepsis is disproportionately higher in low- and middle-income countries. We aimed to determine the prevalence of sepsis, bacteriological profile of causes, antimicrobial susceptibility patterns, and in-hospital outcomes among adult medical emergencies presenting to Kiruddu National Referral Hospital in Kampala, Uganda.

**Methods:** We conducted a prospective cohort study between December 2018 and July 2019 in which patients presenting to the medical emergency ward with sepsis were consecutively enrolled; blood was drawn for aerobic blood cultures, antimicrobial susceptibility patterns were determined and patients were followed up for in-hospital outcomes.

**Results:** Of 1,657 patients screened during the study period, 243(14.7%) had sepsis, the median age was 45 years (IQR 32,65) and the majority were female (55.6%). Among patients with sepsis; 46 (18.9%) had positive aerobic blood cultures. *Staphylococcus aureus w*as the most common isolate (31/46, 67.4%) with a predominance of Methicillin-Resistant *Staphylococcus aureus* (MRSA) (20/31, 64.5%). Of the 243 patients followed up, 143 (58.9%) died in hospital with an average length of stay of 4.9 days (SD 5.5) for those who died and 10.2 (SD 7.6) for those that were discharged alive. None of the patients was admitted to ICU.

**Conclusions:** Sepsis was common (14.7%) among adult medical emergencies and it was associated with a high in-hospital mortality rate (58.9%). Positive blood cultures were predominantly *Staphylococcus aureus* and nearly a third of these were Methicillin-Resistant.

## Introduction

Sepsis occurs when the host immune response to infection is dysregulated resulting in life-threatening organ dysfunction (2). It is a common condition among hospitalized patients with a global estimate of 48.9 million incident cases reported in 2017, resulting in about 11.8 million deaths (3). Low- and middle-income countries (LMICs) are disproportionately affected by sepsis due to the higher prevalence of predisposing conditions such as malnutrition and HIV (4). Mortality from sepsis is higher in LMICs than in developed countries. High-income countries have adopted the current guidelines for sepsis management (1) with resultant improved outcomes including reduced mortality. However, most LMICs lack adequate resources to consistently follow these guidelines (5). Timely initiation of appropriate antimicrobials improves outcomes among patients with sepsis. The choice of empirical antimicrobials is dependent on prevailing local bacteriological and antimicrobial susceptibility profiles (6). In Uganda, this remains a major challenge as is the case in many LMICs where the capacity for identifying specific etiologies and antimicrobial susceptibility patterns is limited (6). This is especially important because of the increasing prevalence of microbial resistance to the most commonly available antibiotics in these settings including Uganda (7).

There is therefore a need for continuous surveillance of sepsis specifically to determine the most prevalent pathogens and their antimicrobial susceptibility patterns. This study sought to determine the prevalence of sepsis, bacteriological profiles, antimicrobial susceptibility patterns, and in-hospital outcomes of patients admitted to Kiruddu national referral hospital in Kampala, Uganda.

## Materials and Methods

### Study design and setting

A prospective observational cohort study was conducted from December 2018 to July 2019 among patients presenting to the medical emergency ward of Kiruddu national referral hospital during the study period. Kiruddu Hospital, located on Buziga Hill, Makindye Division in Kampala is approximately 10 kilometers southeast of the central business district of Kampala. The medical emergency ward has a 25-bed capacity and receives approximately 976 patients per month. The unit acts as a national referral emergency unit caring for adult non-surgical patients. It is run by different cadres of doctors and nurses and has access to the main hospital laboratory and radiology departments for investigations. Patients received in the unit are assessed, stabilized, and then sent to one of the various sub-specialty medical wards based on the presumptive diagnosis. During the study period, the hospital had no facilities for carrying out culture and antimicrobial susceptibility testing and no formal protocol for sepsis identification or management was in place at the unit.

We assessed patients who presented to the unit during the study period for suspected sepsis. Sepsis was defined as the presence of a suspected infection plus two or more clinical proxies for organ dysfunction including oxygen saturation (SPO2) <90%, estimated urine output in the preceding 24 hours <400mls, jaundice, petechiae &/or ecchymoses, Glasgow Coma Scale (GCS) less than 15, systolic blood pressure (SBP)<90mmhg and absent bowel sounds. All adults aged 18 years and above who met the above study definition for sepsis and consented to participate were enrolled after providing written informed consent. Consent was obtained from the next of kin of very ill patients.

### Ethics approval and consent to participate

We obtained approval from the Makerere University School of Medicine Research and Ethics Committee (SOMREC). We also obtained verbal consent before screening all patients and written informed consent before enrollment into the study.

### Data collection and study procedures

The sample size was calculated at 250 and the convenience sampling method was used to assess all patients who presented to the emergency ward. After an initial assessment by the primary healthcare team, we assessed patients for sepsis using the study definition based on an adaptation of the sepsis management recommendations for low-income settings published in 2012 (8). A study questionnaire was administered to either the participant or attendant for very ill patients to collect data on the patient’s history including the sociodemographic and medical history and history of antimicrobial use in the past 30 days. A physical examination was performed at enrolment to identify the possible site(s) of infection, vital parameters to determine the qSOFA score, functional status using the Karnofsky performance scale (KPS), and organ dysfunction. Venous blood was drawn for aerobic blood culture, complete blood count, liver and renal function tests. Other tests included a thick blood smear for malaria or malaria rapid diagnostic test and HIV test (9). Treatment including antimicrobials and fluids was initiated by the attending clinicians and documented by the study team.

### Blood culture and antimicrobial susceptibility testing

Eight milliliters of blood were drawn aseptically and inoculated into an aerobic blood culture bottle (BD BACTEC). The blood samples were transported at room temperature to MBN laboratory within 8 hours of collection, where they were inserted into a blood culture machine (BD BACTEC FX40) immediately to last a protocol length of 5 days. Any growth was detected automatically by the machine and the following procedures were performed; gram-staining and sub-culture on agar media, identification of colonies that grew on the agar, and antibiotic susceptibility testing using the Kirby Bauer disk diffusion method. The susceptibility testing and interpretation were based on the Clinical Laboratory standard institute (CLSI, 2018). All test results were availed to the primary team managing the patients as soon as they were ready. The MBN laboratory participates in an external quality assurance program under the Oneworld Accuracy system.

### Data processing and analysis

All complete data were entered into the computer using a data entry template developed using Epidata version 3.1 software packages with in-built quality control checks. The final data was cleaned, frozen, backed up, and exported to STATA version 14.0 for analysis. Sepsis prevalence was calculated as the proportion of patients with sepsis out of those assessed during the study period. The outcomes were described as proportions for categorical data, means with standard deviations for normally distributed numerical data, and median with interquartile ranges for skewed data. The proportion of aerobic blood culture samples with microbial growth was calculated as a percentage and the isolates were described. The antimicrobial susceptibility patterns (including multidrug resistance) of organisms isolated from blood culture were described.

## Results

Out of the 1647 patients who presented to the medical emergency ward during the study period; 243 fulfilled the study definition for sepsis. Of these, 135 (55.6%) were female and the majority (169 (69.5%) were 60 years or less with a median age of 45 years (IQR=32, 65).

The prevalence of sepsis was 14.7% (243/1647). Fever, confusion, and cough with abnormal lung auscultation were the most common findings at initial assessment, present in 163 (67%), 151(62%), and 129 (53%) respectively. The most common markers of organ dysfunction were GCS <15 and low oxygen saturation (SPO2<90) present in 176 (72%) and 187 (77%) respectively. The majority of patients (53.5%) had used antimicrobials in the preceding 30 days, 86 (35.4%) were HIV positive and 42% had a Karnofsky score of less than 20%.

### In-hospital outcomes

Of the 243 patients with sepsis, 143 (58.9%) died after an average length of hospital stay of 4.9 days. The median duration of hospital stay was longer among those who survived than those who died (7(5,13) vs 3(1,6)) days. None of the patients got transferred to an intensive care unit (ICU) during the study period. Patients with a Glasgow coma scale (GCS) less than 15 at admission were more likely to die in the hospital compared to those with a GCS of 15 (p=0.001) (Table 2).

**Table 1:** Demographic characteristics

| Characteristic | Number<br>243 | Percent<br>100 |
| --- | --- | --- |
| Age: median (IQR) | 45 (32,65) |  |
| Age |  |  |
| Less than or equal to 60 | 169 | 69.6 |
| 61 or above | 74 | 30.4 |
| Sex |  |  |
| Male | 108 | 44.4 |
| Female | 135 | 55.6 |
| Marital Status*7 |  |  |
| Single | 55 | 22.6 |
| Married/Living with a partner | 137 | 56.4 |
| Separated | 51 | 21.0 |
| Education**3 |  |  |
| No formal education | 48 | 20.0 |
| Primary | 88 | 36.7 |
| Secondary | 78 | 32.5 |
| Vocational | 15 | 6.3 |
| University | 11 | 4.6 |
| Active alcohol use***2 |  |  |
| No | 178 | 74.8 |
| Yes | 60 | 25.2 |
| Missing data: **3, ***5 |  |  |

**Table 2:**
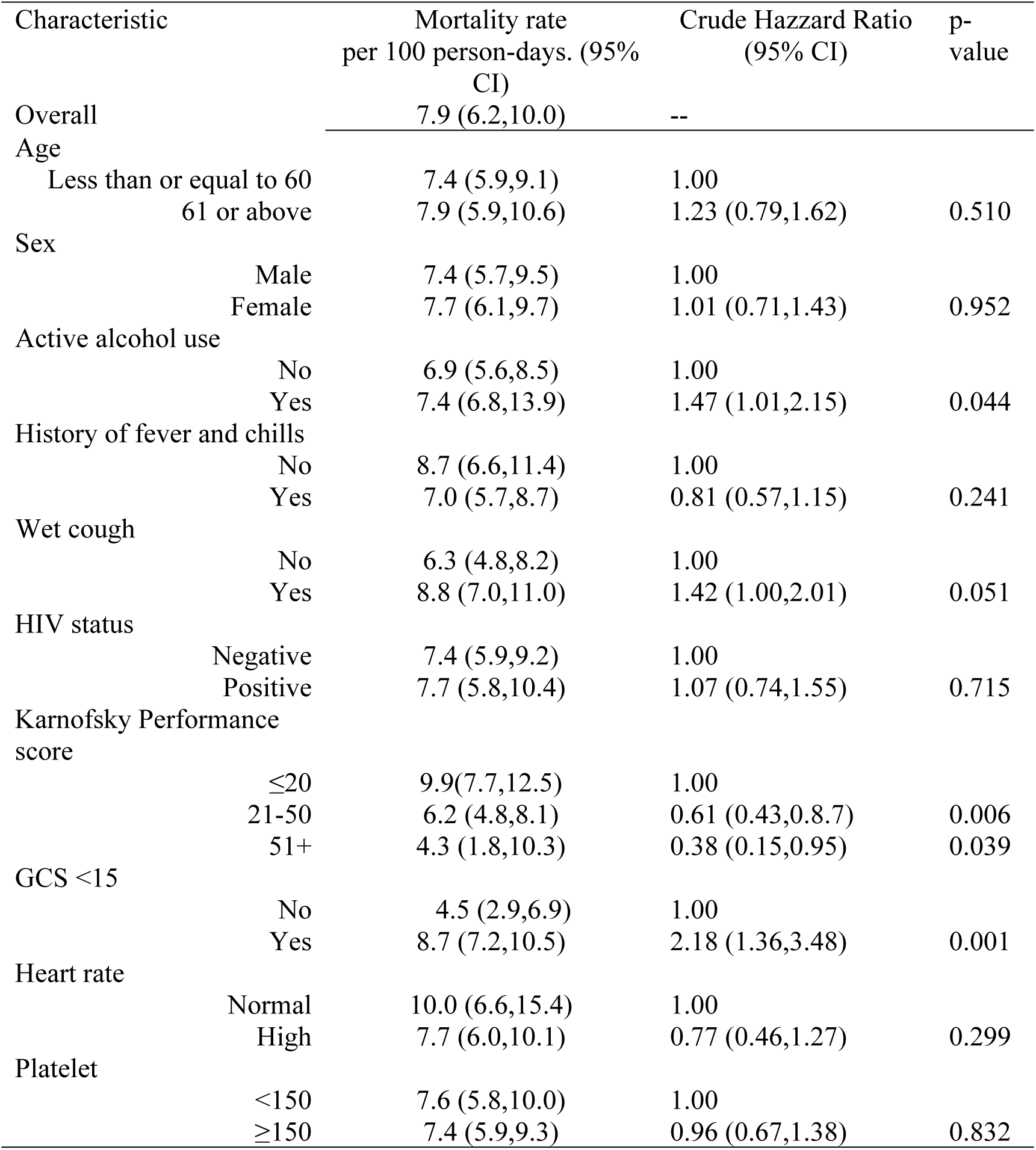
Factors associated with mortality

| Characteristic | Mortality rate<br>per 100 person-days. (95%<br>CI) | Crude Hazzard Ratio<br>(95% CI) | p-<br>value |
| --- | --- | --- | --- |
| Overall | 7.9 (6.2,10.0) | -- |  |
| Age |  |  |  |
| Less than or equal to 60 | 7.4 (5.9,9.1) | 1.00 |  |
| 61 or above | 7.9 (5.9,10.6) | 1.23 (0.79,1.62) | 0.510 |
| Sex |  |  |  |
| Male | 7.4 (5.7,9.5) | 1.00 |  |
| Female | 7.7 (6.1,9.7) | 1.01 (0.71,1.43) | 0.952 |
| Active alcohol use |  |  |  |
| No | 6.9 (5.6,8.5) | 1.00 |  |
| Yes | 7.4 (6.8,13.9) | 1.47 (1.01,2.15) | 0.044 |
| History of fever and chills |  |  |  |
| No | 8.7 (6.6,11.4) | 1.00 |  |
| Yes | 7.0 (5.7,8.7) | 0.81 (0.57,1.15) | 0.241 |
| Wet cough |  |  |  |
| No | 6.3 (4.8,8.2) | 1.00 |  |
| Yes | 8.8 (7.0,11.0) | 1.42 (1.00,2.01) | 0.051 |
| HIV status |  |  |  |
| Negative | 7.4 (5.9,9.2) | 1.00 |  |
| Positive | 7.7 (5.8,10.4) | 1.07 (0.74,1.55) | 0.715 |
| Karnofsky Performance<br>score |  |  |  |
| ≤20 | 9.9(7.7,12.5) | 1.00 |  |
| 21-50 | 6.2 (4.8,8.1) | 0.61 (0.43,0.8.7) | 0.006 |
| 51+ | 4.3 (1.8,10.3) | 0.38 (0.15,0.95) | 0.039 |
| GCS <15 |  |  |  |
| No | 4.5 (2.9,6.9) | 1.00 |  |
| Yes | 8.7 (7.2,10.5) | 2.18 (1.36,3.48) | 0.001 |
| Heart rate |  |  |  |
| Normal | 10.0 (6.6,15.4) | 1.00 |  |
| High | 7.7 (6.0,10.1) | 0.77 (0.46,1.27) | 0.299 |
| Platelet |  |  |  |
| <150 | 7.6 (5.8,10.0) | 1.00 |  |
| ≥150 | 7.4 (5.9,9.3) | 0.96 (0.67,1.38) | 0.832 |

### Antimicrobial susceptibility patterns

The aerobic blood culture positivity rate was 18.9% (46/243) of all blood culture samples. The most commonly isolated organism was *Staphylococcus aureus (S*.*aureus)* with 31 isolates.

Of the *S*.*aureus* isolates, 20, (64.5%) were Methicillin-resistant (MRSA), 31, 100% were resistant to penicillin and 100% were sensitive to vancomycin. Of the 9 Enterobacteriaceae isolates, 8 were resistant to amoxicillin-clavulanic acid, all were resistant to ampicillin and more than half (5) were resistant to ceftriaxone while all were sensitive to amikacin and Imipenem as detailed in table 4 below.

**Table 3.**
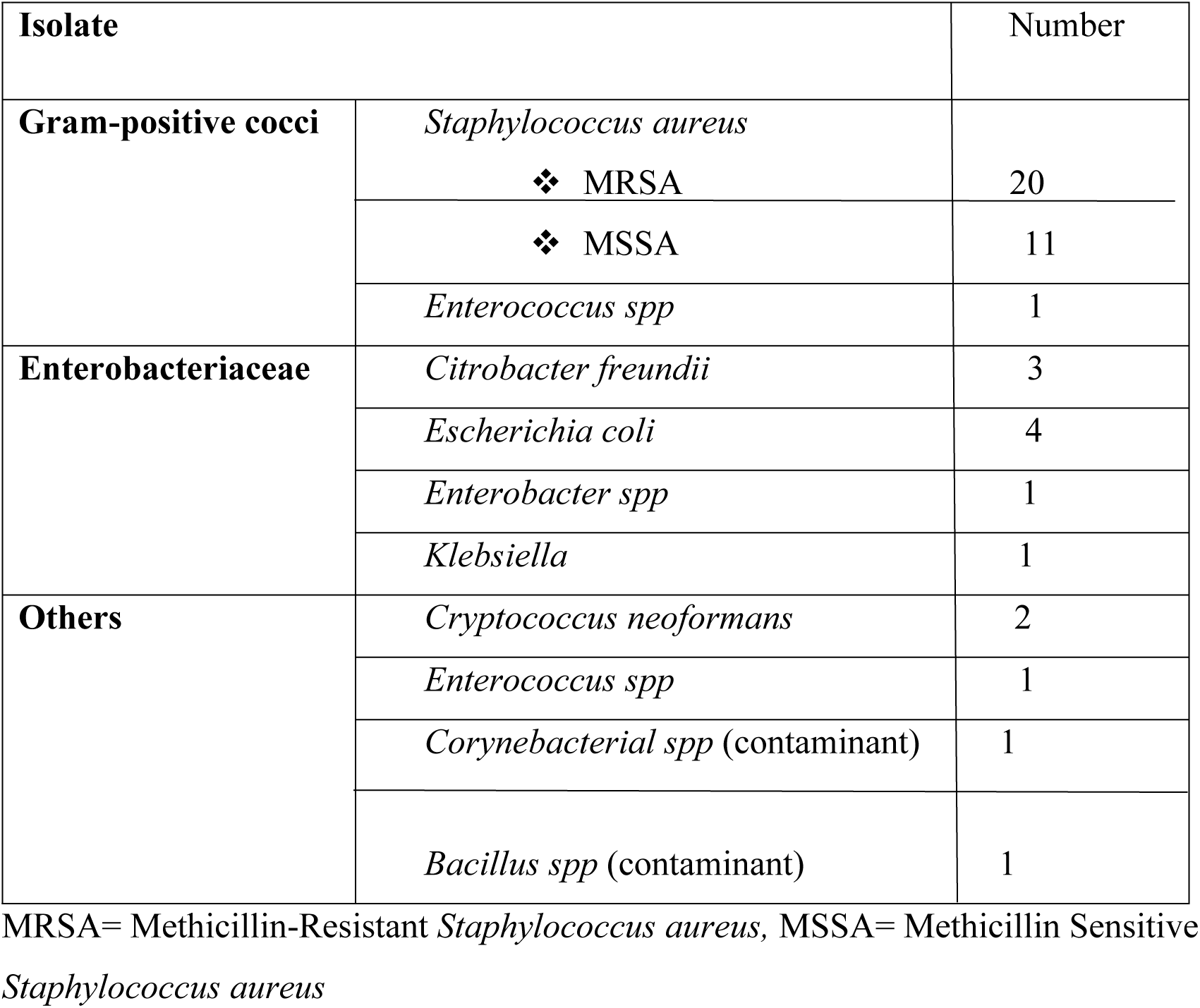
bacterial profile

| Isolate |  | Number |
| --- | --- | --- |
| Gram-positive cocci | *Staphylococcus aureus* |  |
|  | ❖ MRSA | 20 |
|  | ❖ MSSA | 11 |
|  | *Enterococcus spp* | 1 |
| Enterobacteriaceae | *Citrobacter freundii* | 3 |
|  | *Escherichia coli* | 4 |
|  | *Enterobacter spp* | 1 |
|  | *Klebsiella* | 1 |
| Others | *Cryptococcus neoformans* | 2 |
|  | *Enterococcus spp* | 1 |
|  | *Corynebacterial spp* (contaminant) | 1 |
|  | *Bacillus spp* (contaminant) | 1 |

MRSA= Methicillin-Resistant *Staphylococcus aureus*, MSSA= Methicillin Sensitive *Staphylococcus aureus*

**Table 4.** % of Antibiotic resistance among the pathogenic bacterial isolates (N=41)

| **Antibiotic** | ***S.aureus***<br>**(n=31)** | ***Enterococcus***<br>**spp.(n=1)** | ***E. coli***<br>**(=4)** | ***Citrobacter***<br>**spp. (n=3)** | ***Enterobacter***<br>**spp (n=1)** | ***Klebsiella***<br>***pneumoniae***<br>**(n=1)** |
| --- | --- | --- | --- | --- | --- | --- |
| Amikacin | N/T | N/T | 0% | 0% | 0% | 0% |
| Ciprofloxacin | 61% | 0% | 50% | 66% | 0% | 100% |
| Gentamycin | 35% | N/T | 100% | 0% | 100% | 100% |
| Tetracycline | 67% | 100% | N/T | N/T | N/T | N/T |
| Cotrimoxazole | 97% | N/T | 100% | 66% | 100% | 100% |
| Penicillin | 100% | 0% | N/T | N/T | N/T | N/T |
| amoxiclav | N/T | N/T | 75% | 100% | 100% | 100% |
| Ampicillin | N/T | N/T | 100% | 100% | 100% | 100% |
| Ceftriaxone | N/T | N/T | 100% | 66% | 100% | 100% |
| Ceftazidime | N/T | N/T | 50% | 33% | 100% | 0% |
| Ceftaz-clav | N/T | N/T | 33% | 33% | 0% | N/T |
| Cefuroxime | N/T | N/T | 100% | 66% | 100% | 100% |
| CAF | 42% | 0% | 75% | 0% | 100% | 100% |
| Clindamycin | 42% | N/T | N/T | N/T | N/T | N/T |
| erythromycin | 71% | 0% | N/T | N/T | N/T | N/T |
| Oxacillin | 64% | N/T | N/T | N/T | N/T | N/T |
| Imipenem | N/T | N/T | 0% | 0% | 0% | 0% |
| vancomycin | 0% | 0% | N/T | N/T | N/T | N/T |

N/T= Not Tested, CAF=Chloramphenicol, Ceftaz- clav= Ceftazidime- Clavulanic acid

Of the Enterobacteriaceae, two Isolates were producers of Extended Spectrum Beta-lactamases (ESBL).

## Discussion

We aimed to determine the prevalence of sepsis, bacteriological profiles, antimicrobial susceptibility patterns, and in-hospital outcomes of patients admitted with sepsis at Kiruddu national referral hospital in Kampala, Uganda. The prevalence of sepsis was high at 14.7%. This is higher than the prevalence reported in the few studies that assessed sepsis outside the ICU. For example, a study in Brazil done about 10 years ago found a sepsis occurrence rate of 6.4% in the emergency department (10). A more recent study assessing the point prevalence of sepsis in general medical/emergency wards in Wales found a prevalence of 5.5% (11). The differences may be due to differences in criteria used to define sepsis. The different criteria for sepsis diagnosis highlight the challenges faced in the identification of sepsis, especially in low-income settings such as Uganda. This is due to the lack of specific sepsis signs and symptoms without a gold standard test for diagnosis. The relatively high prevalence found in this study is also in agreement with the consensus that there is a higher sepsis burden in low-income settings (12).

### In-hospital outcomes

We found a mortality rate of 57.8% within an average duration of hospital stay of 4.9 days. The median duration of hospital stay was longer among those who survived than those who died (7(5,13) vs 3(1,6)), meaning patients died early on during hospitalization. This mortality rate is higher than that found by previous studies in Uganda. Jacob and colleagues found an in-hospital mortality rate of 23.7% and an overall mortality rate of 43% up to 30days post-discharge (Jacob et al., 2009). Another study from western Uganda reported in-hospital mortality of 30% (13). The differences in mortality rates could be explained by the severity of illness of patients in this study where over 42% had a Karnofsky score of ≤ 20%. None of the patients in this study was admitted to the ICU within 30 days of in-hospital follow-up. This lack of intensive care management plus the presence of drug-resistant bacteria as evidenced by 8 out of the 9 *Enterobacteriaceae spp* isolates resistant to ceftriaxone, most likely contributed to the high mortality rates in this study. Ceftriaxone is the most common antibiotic prescribed empirically in the medical emergency ward. In addition, 64.5% of the *S*.*aureus* isolates were MRSA yet vancomycin was not readily available. Studies from other settings found similarly high mortality rates; for example, one recent study from India reported in-hospital mortality of 63.6% (14). Overall, this indicates that sepsis contributes significantly to mortality in hospitals worldwide.

### Aerobic blood culture and antimicrobial susceptibility patterns

The aerobic blood culture growth rate was 18.9% in this study with *Staphylococcus aureus (S. aureus*) as the most frequently isolated pathogen (31 of the 46 isolates). More than half of the S. aureus isolated were MRSA. The relatively low culture positivity rate may have been because more than half (53.5%) of the patients had been treated with antimicrobials within the 30 days before admission. Irrational use of antimicrobials is rampant in many low-income settings including in Uganda. A study by Mukonzo et al (2013) found that 41% of all antibiotic sales at community pharmacies were over the counter and not accompanied by a prescription (15). Also, one blood culture set was obtained yet two or more might have increased the yield. However, other studies in Uganda where aerobic blood cultures were performed reported similar rates of culture positivity. For example, a recent study summarizing antimicrobial susceptibility patterns reported culture growth in 14% of the samples. This study also reported *S. aureus* as the most frequently identified growth and 32% of these were methicillin-resistant (16). An earlier study reported a similar rate of culture-positive sepsis of 20% and more than half of the patients (57.7%) had used antimicrobials before admission., but the commonest isolate was non-Typhoidal Salmonella species(17). Low blood culture yields have also been reported in many studies outside the Ugandan setting. The high rate of drug-resistant pathogens identified provides evidence for the growing problem of antimicrobial resistance in Uganda. This is likely contributed to by the inadequate anti-microbial stewardship in the country.

## Conclusion

The prevalence of sepsis among patients presenting to the emergency unit of Kiruddu National Referral Hospital was high at 14.7% and associated with a very high in-hospital mortality rate of 58.9%. None of the patients were admitted to the intensive care unit during the study period. Culture negative sepsis was common. Among patients with culture-positive sepsis, drug-resistant bacteria such as MRSA were highly prevalent.

## Data Availability

data are not currently available in a public repository but can be availed up on request from the corresponding author

## Acknowledgments

We thank the Kiruddu national referral hospital medical emergency ward staff for their assistance during data collection and the study participants for accepting to take part in the study.

## References

1. Singer M, Deutschman CS, Seymour C, Shankar-Hari M, Annane D, Bauer M, et al. The third international consensus definition for sepsis and septic shock (sepsis-3). JAMA - Journal of the American Medical Association. 2016.

2. Rhodes A, Bs MB, Co-chair R, Evans LE, Co-chair F, Alhazzani W, et al. Surviving Sepsis Campaign : International Guidelines for Management of Sepsis and Septic Shock : 2016. 2017.

3. Rudd KE, Johnson SC, Agesa KM, Shackelford KA, Tsoi D, Kievlan DR, et al. Global, regional, and national sepsis incidence and mortality, 1990–2017: analysis for the Global Burden of Disease Study. Lancet. 2020;

4. Jacob ST, Moore CC, Banura P, Pinkerton R, Meya D, Opendi P, et al. Severe sepsis in two Ugandan hospitals: A prospective observational study of management and outcomes in a predominantly HIV-1 infected population. PLoS One. 2009;

5. Jacob ST, West TE, Banura P. Fitting a square peg into a round hole : are the current Surviving Sepsis Campaign guidelines feasible for Africa ? 2011;1–3.

6. Jacob ST, Banura P, Baeten JM, Moore CC, Meya D, Nakiyingi L, et al. prospective intervention study. 2013;40(7):2050–8.

7. Blondeau JM, Vaughan D. A review of antimicrobial resistance in Canada. Can J Microbiol. 2000;46(10):867–77.

8. Dünser MW, Festic E, Dondorp A, Kissoon N, Ganbat T, Kwizera A, et al. Recommendations for sepsis management in resource-limited settings. Intensive Care Med. 2012;38(4):557–74.

9. Ministry of Health Uganda. Consolidated Guidelines on the prevention and treatment of HIV in Uganda. 2018;(August):1–200.

10. Rezende E, Silva Junior JM, Isola AM, Campos EV, Amendola CP, Almeida SL. Epidemiology of severe sepsis in the emergency department and difficulties in the initial assistance. Clinics [Internet]. 2008;63(4):457–64. Available from: http://www.scielo.br/scielo.php?script=sci_arttext&pid=S1807-59322008000400008&lng=en&nrm=iso&tlng=en

11. Szakmany T, Lundin RM, Sharif B, Ellis G, Morgan P, Kopczynska M, et al. Sepsis prevalence and outcome on the general wards and emergency departments in Wales: Results of a multi-centre, observational, point prevalence study. PLoS One. 2016;11(12):1–12.

12. Rudd KE, Kissoon N, Limmathurotsakul D, Bory S, Mutahunga B, Seymour CW, et al. The global burden of sepsis: barriers and potential solutions. Crit Care [Internet]. 2018 Sep 23;22(1):232. Available from: https://www.ncbi.nlm.nih.gov/pubmed/30243300

13. Ssekitoleko R, Pinkerton R, Muhindo R, Bhagani S, Moore CC. Aggregate Evaluable Organ Dysfunction Predicts In-Hospital Mortality from Sepsis in Uganda. 2011;85(4):697–702.

14. Chatterjee S, Bhattacharya M, Todi SK. Epidemiology of Adult-population Sepsis in India: A Single Center 5 Year Experience. Indian J Crit Care Med [Internet]. 2017 Sep;21(9):573–7. Available from: https://www.ncbi.nlm.nih.gov/pubmed/28970656

15. Mukonzo JK, Namuwenge PM, Okure G, Mwesige B, Namusisi OK, Mukanga D. Over-the-counter suboptimal dispensing of antibiotics in Uganda. J Multidiscip Healthc [Internet]. 2013 Aug 20;6:303–10. Available from: https://pubmed.ncbi.nlm.nih.gov/23990728

16. Kajumbula H, Fujita AW, Mbabazi O, Najjuka C, Izale C, Akampurira A, et al. Antimicrobial drug resistance in blood culture isolates at a tertiary hospital, Uganda. Emerg Infect Dis. 2018;24(1):174–5.

17. Jacob ST, Moore CC, Banura P, Pinkerton R, Meya D, Reynolds SJ, et al. Severe Sepsis in Two Ugandan Hospitals : a Prospective Observational Study of Management and Outcomes in a Predominantly HIV-1 Infected Population. 2009;4(11).

